# Predicting prolonged mechanical ventilation after endovascular treatment for acute vertebrobasilar artery occlusion

**DOI:** 10.1101/2023.12.05.23299562

**Authors:** Qiankun Cai, Minying Hong, Yingjie Xu, Shuai Zhang, Zhixin Huang, Pengfei Xu, Xianjun Huang, Shufen Liu, Zhengping Huang, Lichao Ye, Jixing Chen, Hongyu Huang, Feng Zheng, Wen Sun

## Abstract

**Background:** Patients with acute vertebrobasilar artery occlusion (VBAO) after endovascular treatment (EVT) frequently require mechanical ventilation (MV). This study aims to develop and validate a predictive score for prolonged mechanical ventilation (PMV) in patients with VBAO after EVT.

**Methods:** The derivation cohort prospectively recruited acute VBAO patients undergoing EVT from four comprehensive stroke centers (CSCs) in China. PMV was defined as continuous MV for ≥96 hours. Two multivariable logistic regressions were performed to predict predictive models based on whether adding radiographic results at follow-up. The performance of two models was evaluated for discrimination, calibration, and clinical utility. A validation cohort of 444 patients from acute Posterior circulation ischemic Stroke registry was enrolled to validate two models.

**Results:** The derivation cohort consisted of 498 patients from four CSCs, among whom 204 (41.0%) required PMV. Reperfusion status, National Institutes of Health Stroke Scale scores, collateral circulation status, history of atrial fibrillation, fasting blood glucose, circulation insufficiency, the presence of malignant cerebellar edema, and symptomatic intracranial hemorrhage at follow-up imaging were found to be independent predictors of PMV in multivariable logistic regression. These variables were combined to create the basic and radiographic PAIRS scores. The basic PAIRS and radiographic PAIRS scores demonstrated adequate discrimination in derivation cohort (C-index, 0.77 [95%CI 0.73 to 0.81] and 0.80 [95%CI 0.77 to 0.84]), as well as the validation cohort (C-index, 0.70 [95%CI 0.65 to 0.75] and 0.75 [95%CI 0.70 to 0.80]), and had well-fitted calibration plot for both cohorts. Decision curve analysis showed that both PAIRS scores had satisfied net benefit in predicting PMV across various thresholds.

**Conclusions:** The PAIRS scores provide adequate accurate predictions of PMV in VBAO patients after EVT. These findings can assist patient families and caregivers in better understanding the treatment trajectory.

## INTRODUCTION

Vertebrobasilar artery occlusion (VBAO) is a rare but severe form of ischemic stroke, accounting for 1% of all strokes and 5% of large vessel occlusions (LVOs).^1,2^ Unlike anterior circulation LVOs, VBAO often results in the need for invasive mechanical ventilation (MV) due to its impact on brain areas responsible for consciousness, breathing, and swallowing. This increased risk of respiratory failure contributes to higher rates of disability and mortality associated with VBAO.^3^ Prior research indicates an alarming 88% mortality rate among VBAO patients requiring MV.^4^ Recent clinical trials have demonstrated that combining endovascular treatment (EVT) with optimal medical care improves functional outcomes at 90 days compared to medical care alone.^5,6^ In a comprehensive, nationally representative cohort of hospitalized acute ischemic stroke patients, approximately 16.7% of those undergoing EVT necessitated prolonged mechanical ventilation (PMV), defined as continuous invasive MV for ≥96 hours. This group had a significantly higher in-hospital mortality rate compared to those who did not require PMV (32.1% vs 8.2%). It’s important to note that the study primarily focused on patients with anterior circulation LVOs.^7^

Given the distinct divergence in MV requirements and outcomes between anterior and posterior circulation LVOs patients, coupled with the encouraging advancements in EVT for VBAO, there is a conspicuous absence of a predictive model for identifying VBAO patients at risk of requiring PMV post-EVT. This study endeavors to develop and validate a risk model, termed the **P**rolonged mech**A**nical vent**I**lation after endovascular treatment for acute verteb**R**obasilar artery occlu**S**ion (PAIRS) score.

## METHODS

The data that support the finds of this study are available from the corresponding author on reasonable request.

### Derivation cohort

We enrolled VBAO patients who underwent EVT at four comprehensive stroke centers (First Affiliated Hospital of University of Science and Technology of China [USTC], Guangdong second Provincial General Hospital, the Second Affiliated Hospital of Fujian Medical University, The Affiliated hospital of Yangzhou University) between January 2019 and June 2022. Inclusion criteria were: (1) age ≥18 years; (2) imaging confirmation of acute symptomatic VBAO; and (3) EVT performed within 24 hours of estimated occlusion. Patients with only posterior cerebral artery occlusion treated with EVT and a pre-stroke modified Rankin Scale (mRS) score >2 were excluded. Additionally, we excluded patients without MV information (n=12), those who died within 96 hours while on MV (n=29), and those in whom MV was initiated ≥7 days after VBAO onset (n=30). These stringent criteria were employed to ensure the homogeneity of our study group and the reliability of our findings in the context of prolonged mechanical ventilation post-EVT in VBAO cases. This study was approved by ethics committee of USTC: No 2020-40; Guangdong Second Provincial General Hospital: No 2020-KY-KZ-100-03; the Second Affiliated Hospital of Fujian medical University: No 2020-339; The Affiliated hospital of Yangzhou University: No 2020-153. Written informed consents were obtained from patients or authorized representatives.

### External validation cohort

The acute Posterior circulation ischemic Stroke registry (PERSIST) database is a retrospective registry of patients with acute VBAO who underwent EVT within 24 hours of estimated occlusion time (EOT) between December 2015 and December 2018 at 21 stroke centers in 13 provinces of China (http://www.chictr.org.cn/; unique identifier: ChiCTR2000033211). Details of the PERSIST study have been described previously.^8–10^ In this study we further excluded patients with only posterior cerebral artery occlusion treated with EVT (n=24), those without records of MV information (n=58), those who died within 96 hours while on MV (n=34), and those in whom MV was initiated ≥7 days after VBAO onset (n=29). The PERSIST study was approved by the ethics committee of each participating center.

### Weaning protocol

Generally, all patients were monitored in neurocritical intensive care unit for at least 24 hours post-EVT. Patients using MV were checked daily by physicians at each center according to the predefined weaning criteria: (1) stable cardiovascular status (heart rate[HR] ≤ 140 beats/min, systolic blood pressure[SBP] 90-160 mmHg, and minimal or absence of catecholamine); (2) adequate oxygenation (oxygen saturation measured by pulse oximetry [Spo_2_]≥90%, fractional inspired oxygen tension [Fio_2_] ≤40%, positive end-expiratory pressure ≤8cm H_2_O, respiratory rate ≤35 breaths/min); (3) PaCO_2_ ≤50 mmHg; (4) temperature <38.5℃; (5) pH >7.35. When patients met the criteria above, a spontaneous breathing trial was performed with 30-min T-tube trial or ventilatory support level ≤ 7 to 8 cm H_2_O. Failure of weaning test was defined as the development within 30min of any the following criteria: respiratory rate >35 breaths/min with increased accessory muscle activity, Spo_2_ <90% (on Fio_2_=0.4), HR >140 beats/min, SBP <90 mmHg or >180 mmHg, major dyspnea or agitation.^11^

### Data collection

Patient characteristics, medical history, level of fasting blood glucose (FBG), stroke cause, pre-treatment imaging parameters, treatment-related factors, and clinical examination and radiographic results at follow-up were meticulously gathered. All patients underwent regular clinical examination, including assessment of consciousness using the Glasgow Coma Scale score (GCS), evaluation of stroke severity with the National Institutes of Health Stroke Scale (NIHSS) score, and grading of illness severity at ICU admission using the Simplified Acute Physiology Score (SAPS II)^12^ at admission by each center.

Imaging parameters were collected, including infarction core status, assessed using the posterior circulation Alberta Stroke Program Early CT Score (pc-ASPECTS) on CT or MRI^13^; posterior circulation collateral status, evaluated through the Basilar artery on CT angiography (BATMAN)^14^; and reperfusion status, determined according to modified Thrombolysis in Cerebral Infarction scale on final angiography, with scores of 2b or 3 indicating successful reperfusion^15^. The occlusion site was categorized as either ‘basilar artery + vertebral artery’ or solely ‘basilar artery’ based on specific criteria.^9^ Typically, patients underwent CT or MRI scans within 72 hours post-EVT for radiographic assessment.

Symptomatic intracranial hemorrhage (sICH) was defined as neuroimaging-identified intracranial hemorrhage coupled with ≥4-point increase in NIHSS score.^16^ Malignant cerebellar edema (MCE) was determined using the Jauss scale for neuroradiological mass effect, with a score of ≥ 4 indicating MCE.^17,18^ All neuroimaging data were sent to the core laboratory of USTC and evaluated blindly by two experienced neuroradiologists, with any discrepancies resolved through consensus.

Treatment-related factors such as the choice of first-line treatment, intravenous thrombolysis, time from EOT to puncture, time from puncture to reperfusion, type of anesthesia administered, and any instances of circulation insufficiency (defined as the use of vasoconstrictors like norepinephrine, isoproterenol, and dopamine during hospitalization) were also documented.

### Outcomes

The primary outcome was the development of PMV, which was identified using International Classification of Disease-9th Revision-Clinical Modification (ICD-9 CM) procedural code 96.72 or ICD-10 code Z99.1, defined as continuous invasive MV for ≥ 96 hours.^7,19^

Secondary outcomes included unfavorable outcome (mRS score of 4-6) at 90 days, mortality at 90 days and in-hospital mortality.

### Statistical analysis

Data were presented as the median with interquartile range or numbers with percentages. Univariate analysis was performed using the χ^2^ test for dichotomous variables and Mann-Whitney U test for continuous variables to compare patients with and without PMV in derivation cohort. For the missing variables, multivariate imputation by chained equation was performed using the fully conditional specification approach.^7^ The missing data of study are shown in Table S1.

Two logistic regression analyses were utilized to identify the independent predictors of PMV based on whether further adding radiographic results at follow-up in derivation cohort. Model 1 included variables from patient characteristics, medical history, FBG level, stroke cause, pre-treatment imaging parameters, treatment-related factors, and clinical examination with a *P*-value <0.05 from univariate analyses. Model 2 incorporated variables from model 1 as well as radiographic results at follow-up with a *P*-value <0.05 from univariate analyses. Regression coefficients, odds ratios (OR), and 95% confidence intervals (95% CI) for each variable in the models were calculated.

The PAIRS scores (basic and radiographic) were developed using β coefficients obtained from two logistic regression analyses.^20^ The sum scores for each patient were calculated based on the respective PAIRS score systems. To evaluate prediction accuracy, we used three methods: concordance index (C-index) with 95% CI to measure discrimination, calibration plot to measure model fit, and decision curve analysis to measure clinical utility. Internal validation was performed by generating 1000 bootstrap resamples from the derivation cohort to ensure robustness. External validation was conducted using data from the validation cohort. A clinically useful C-index was considered to be ≥ 0.70.

Category-free net reclassification improvement (NRI) and integrated discrimination improvement (IDI) were used to evaluate whether radiographic PAIRS score compared with basic PAIRS could improve predictive performance.^21^

All statistical analyses were performed with SAS version 9.4 and R version 4.3.1. A two-sided *P* value <0.05 was considered statistically significant. This study is reported as per Transparent Reporting of a Multivariable Prediction Model for Individual Prognosis or Diagnosis guidelines.

## RESULTS

The study included 593 VBAO patients who underwent EVT in the derivation cohort and 609 patients in the validation cohort. Out of these, 498 patients (median age: 66 years, 28.9% female) from the derivation cohort and 444 patients (median age: 66 years, 28.4% female) from the validation cohort were eligible for analysis. Figure 1 presents the flowchart for patient selection, and Table 1 shows the characteristics of patients in both cohorts.

**Figure 1.**
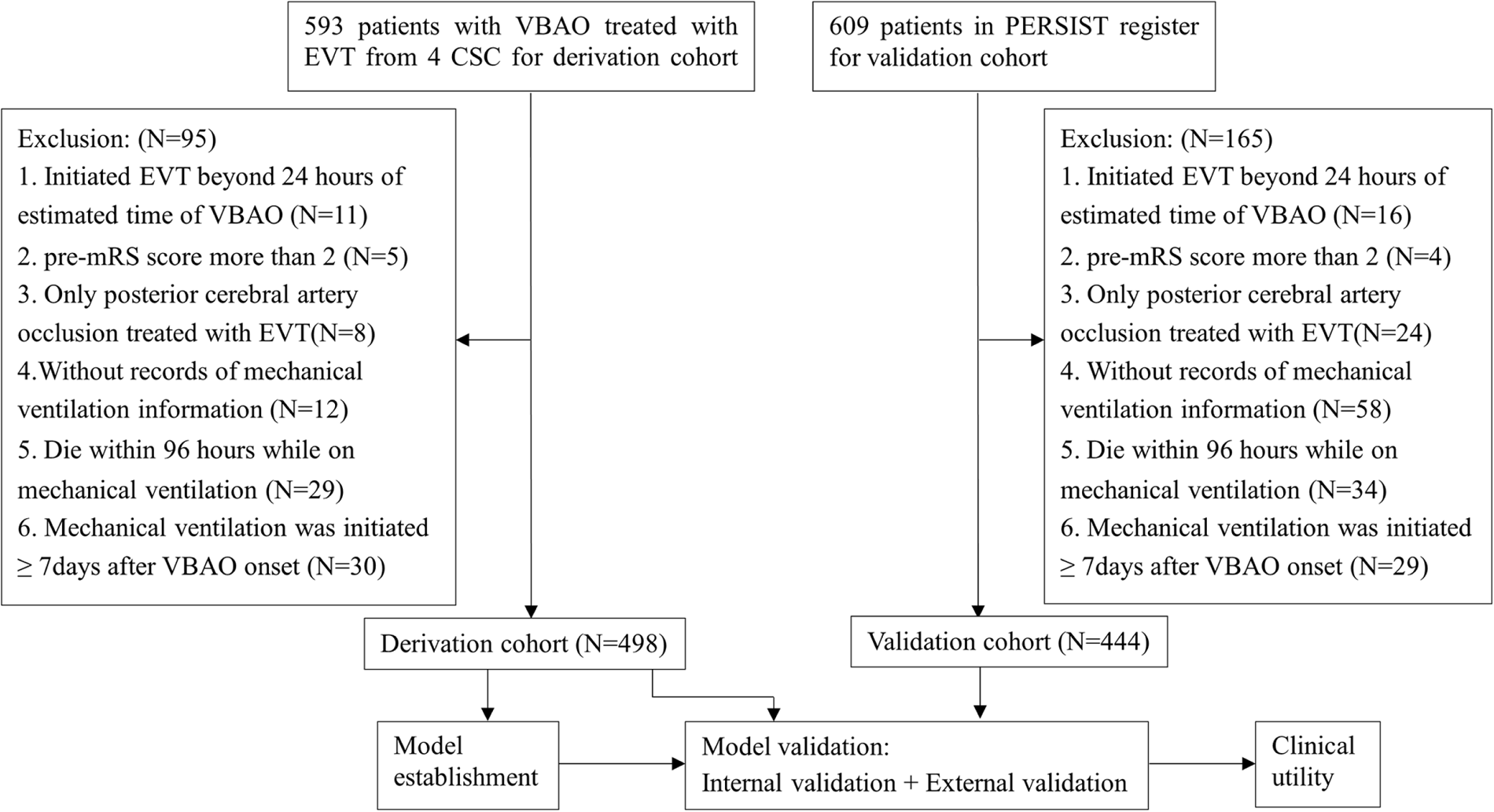
Flow chart of patient inclusion. VBAO, vertebrobasilar artery occlusion; EVT, endovascular treatment; CSC: comprehensive stroke centers; PERSIST, acute Posterior circulation ischemic Stroke registry; mRS: modified Rankin Scale.

**Table 1.** Baseline characteristics of the cohorts.

|  | Derivation cohort | Validation cohort |  |
| --- | --- | --- | --- |
|  | N=498 | N=444 | *P* |
| Age, years | 66(57-74) | 66(57-73) | 0.467 |
| Female | 144(28.9) | 126(28.4) | 0.856 |
| **Medical history** |  |  |  |
| Hypertension | 357(71.7) | 305(68.7) | 0.316 |
| Diabetes mellitus | 125(25.1) | 96(21.6) | 0.208 |
| Hyperlipidemia | 178(35.7) | 156(35.3) | 0.886 |
| Atrial fibrillation | 120(24.1) | 97(21.8) | 0.413 |
| Previous stroke | 118(23.7) | 101(22.9) | 0.774 |
| Coronary artery disease | 52(10.4) | 55(12.4) | 0.348 |
| Current smoking | 147(29.5) | 140(31.5) | 0.503 |
| Drink | 97(19.5) | 101(22.7) | 0.219 |
| Fasting blood glucose, mmol/L | 7.5(6.1-9.8) | 6.9(5.9-8.8) | 0.001 |
| **Clinical examination** |  |  |  |
| Glasgow Coma Scale | 7(6-11) | 9(6-13) | <0.001 |
| NIHSS | 23(14-30) | 22(13-29) | 0.126 |
| SAPS II | 33(27-40) | 32(22-39) | 0.107 |
| Stroke cause |  |  | 0.354 |
| Large artery atherosclerosis | 319(64.1) | 279(62.8) |  |
| Cardioembolism | 107(21.5) | 99(22.3) |  |
| Other | 25(5.0) | 14(3.2) |  |
| Undetermined | 47(9.4) | 52(11.7) |  |
| **Pre-treatment imaging parameters** |  |  |  |
| pc-ASPECTS | 9(8-10) | 8(7-10) | <0.001 |
| BATMAN | 4(2-6) | 6(3-8) | <0.001 |
| Site of occlusion |  |  | 0.005 |
| Basilar artery | 314(63.1) | 318(71.6) |  |
| Basilar artery + vertebral artery | 184(36.9) | 126(28.4) |  |
| **Treatment-related factors** |  |  |  |
| Intravenous thrombolysis | 90(18.1) | 70(15.8) | 0.347 |
| Estimated occlusion to puncture time, minutes | 321(220-466) | 350(211-540) | 0.096 |
| Puncture to reperfusion time, minutes | 109(70-152) | 112(75-150) | 0.681 |
| General anesthesia | 119(23.9) | 182(41.0) | <0.001 |
| First-line treatment |  |  | <0.001 |
| Stent retriever | 330(66.3) | 372(84.5) |  |
| Aspiration | 88(17.7) | 33(7.5) |  |
| Balloon angioplasty or stenting | 68(13.7) | 29(6.6) |  |
| Intra-arterial thrombolysis | 12(2.4) | 6(1.4) |  |
| Rescue therapy | 234(47.0) | 145(32.7) | <0.001 |
| Successful reperfusion | 427(85.7) | 389(87.6) | 0.400 |
| Circulation insufficiency | 127(25.5) | 31(7.0) | <0.001 |
| **Radiographic results at follow-up** |  |  |  |
| sICH | 45(9.0) | 31(7.0) | 0.248 |
| Malignant cerebellar edema | 93(18.7) | 44(9.9) | <0.001 |

Data are present as n (%). Continuous variables are expressed as median (interquartile range). BATMAN, basilar artery on CT angiography; NIHSS, National Institutes of Health Stroke Scale; pc-ASPECTS, posterior circulation-Alberta Stroke Program Early CT score; SAPS II, Simplified Acute Physiology Score; sICH, symptomatic intracranial hemorrhage.

### The primary outcome and secondary outcome

In the derivation cohort, 204 patients (41.0%) required PMV, while in the validation cohort, 143 (32.2%) needed PMV. Patients requiring PMV had significantly higher rates of unfavorable outcomes, mortality at 90 days, and in-hospital mortality compared to those who did not require PMV in both cohorts. Multivariable logistic regressions, after adjusting for confounding variables, demonstrated that PMV was independently associated with unfavorable outcomes, mortality at 90 days, and in-hospital mortality (see Figure S1). The distribution of mRS scores at 90 days based on PMV status in the two cohorts is shown in Figure S2.

### Model development

A comparison of baseline characteristics, medical history, clinical examination, treatment-related factors, and imaging parameters between non-PMV and PMV in derivation cohort is shown in Table 2. Compared with non-PMV patients, patients with PMV were significantly less likely to be female (*P*=0.045), have atrial fibrillation (AF) (*P*=0.007), receive intravenous thrombolysis (*P*=0.019), have good collateral (less BATMAN score, *P*<0.001) and achieve successful recanalization (*P*<0.001); they were more likely to present with severer clinical examination score (a higher NIHSS score, higher SAPS Ⅱ score, and a lower GCS score) and higher FBG level (all *P*<0.001); they had a higher proportion of circulation insufficiency, presence of sICH, and MCE at follow-up imaging (all *P*<0.001).

**Table 2.**
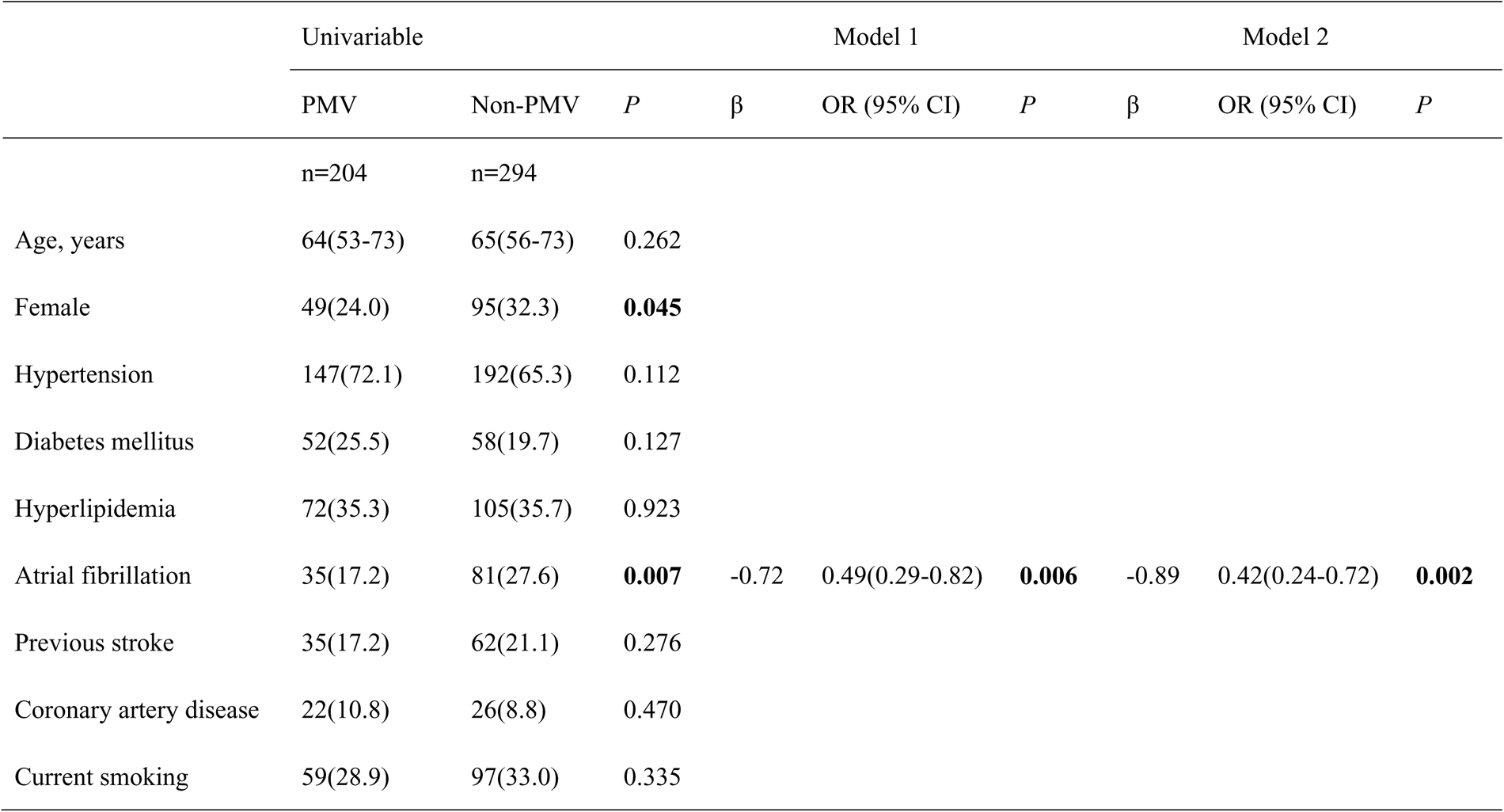
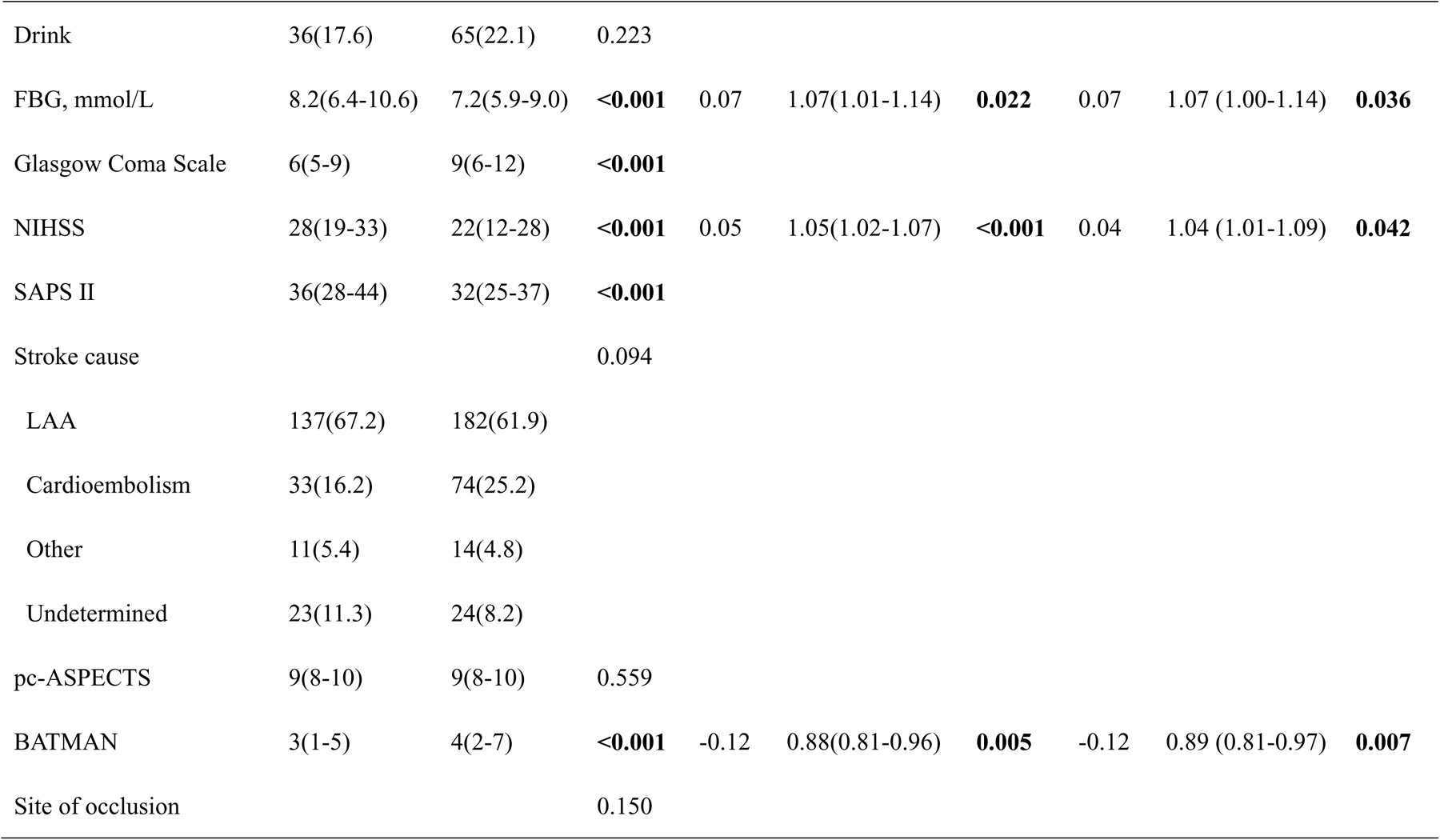
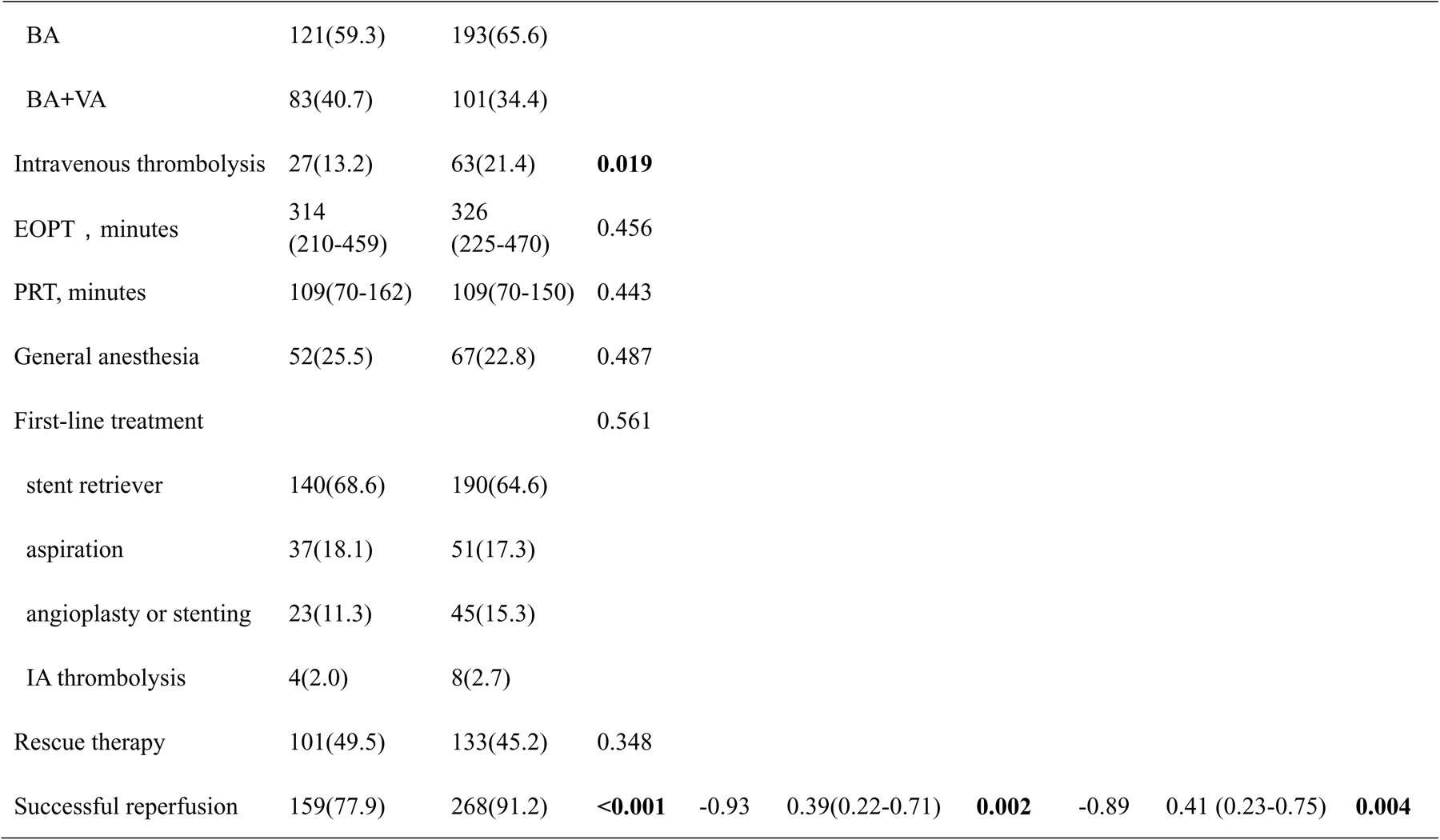
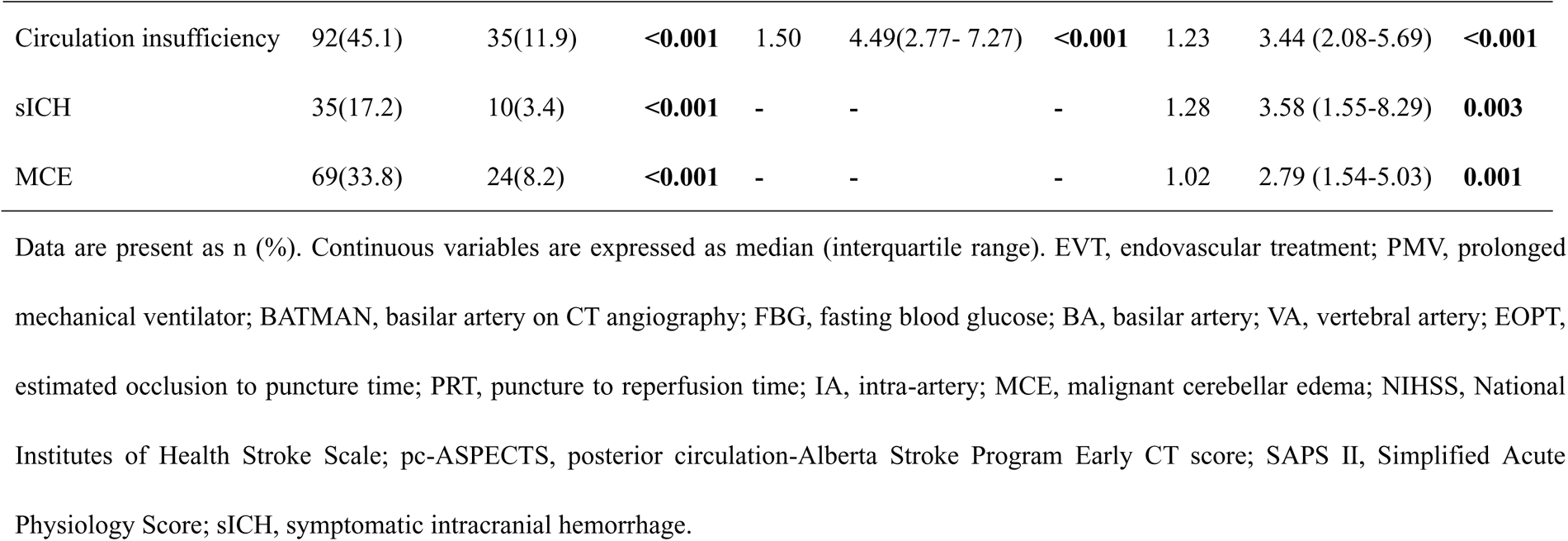
Results of the univariable and multivariable logistic regression for PMV after EVT in the derivation cohort.

In model 1, multivariate logistic analysis found that reperfusion status, posterior circulation collateral status, NIHSS score on admission, FBG level, history of AF, and circulation insufficiency were independently associated with PMV (Table 2). In model 2, the presence of sICH and MCE were also independently associated with PMV (Table 2).

The PAIRS scores were developed based on the β coefficients of individual variables in the two models for predicting PMV. In the derivation cohort, the maximum achievable score was 24 for the basic PAIRS score and 26 for the radiographic PAIRS score (Figure 2A). The C-index for the basic PAIRS score was 0.77 (95% CI 0.73 to 0.81), and for the radiographic PAIRS score, it was 0.80 (95% CI 0.77 to 0.84). Calibration plots in the derivation and internal validation (see Figure S3) demonstrated that the predicted probabilities of PMV were close to the observed PMV.

**Figure 2.**
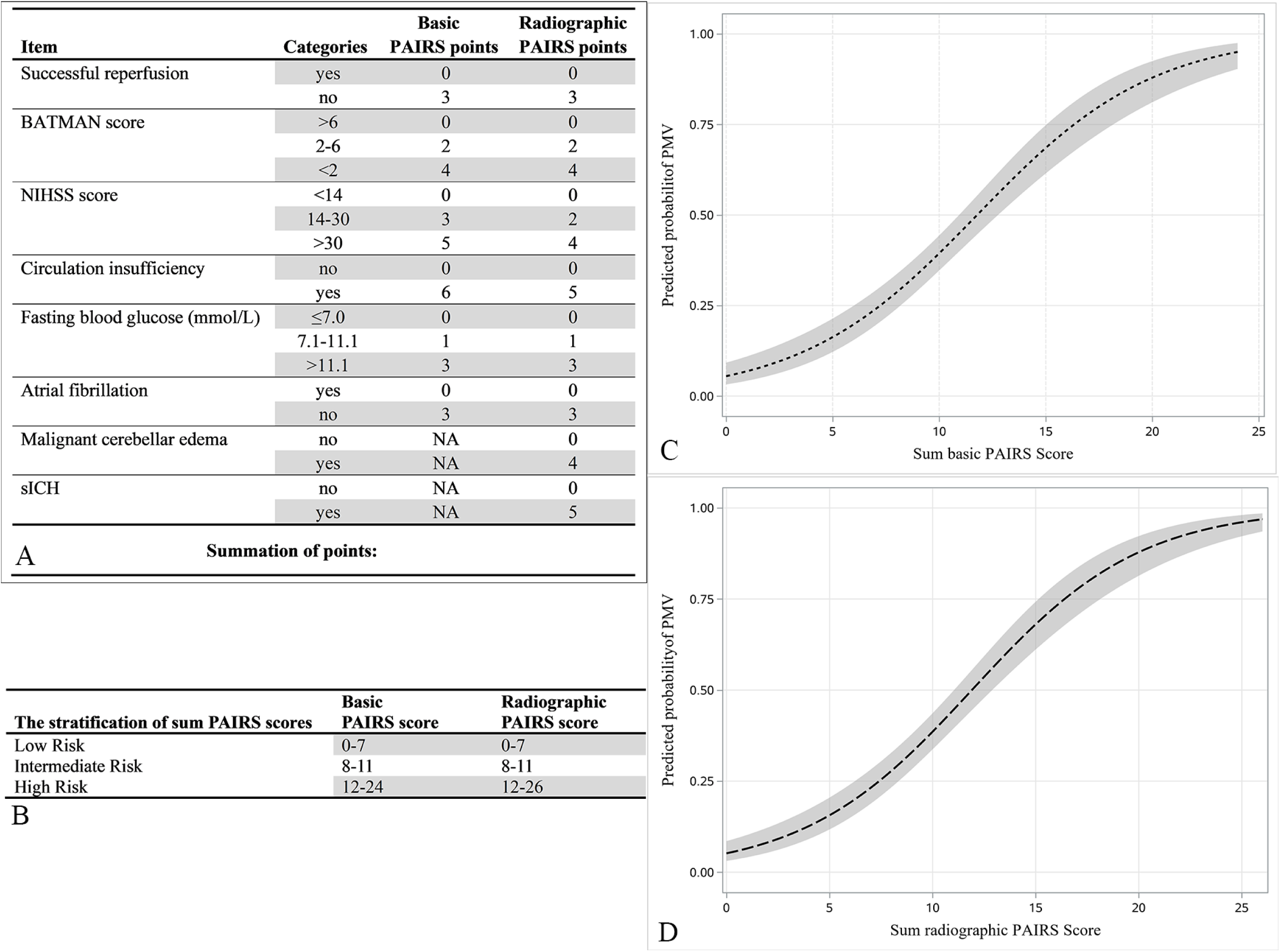
Components of the two PAIRS grading scale (A); The stratification of the two sum PAIRS scores (B); The association between the sum PAIRS score and predicted probability with 95% confidence of PMV (C for basic PAIRS) and (D for radiographic PAIRS). PMV, prolonged mechanical ventilation; BATMAN, basilar artery on CT angiography; NIHSS, National Institutes of Health Stroke Scale. sICH, symptomatic intracranial hemorrhage.

As the sum score values of the two PAIRS scores increased, the predicted probability of PMV also increased gradually (Figure 2C, 2D and Table S2). In the derivation cohort, scores <8 indicated a low risk (0-25%) for PMV, 8-11 points indicated an intermediate risk (26-50%), and ≥12 points indicated a high risk (>50%) for PMV (Figure 2B). The association between three risk stratifications of sum PAIRS score and actual PMV were displayed in Figure S4.

### Model validation

The internal validation yielded a bootstrap-corrected C-index of 0.77 (95% CI 0.75 to 0.78) for the basic PAIRS score and 0.80 (95% CI 0.78 to 0.81) for the radiographic PAIRS score.

In the external validation, the basic PAIRS score had a C-index of 0.70 (95% CI 0.65 to 0.75) and the radiographic PAIRS score had a C-index of 0.75 (95% CI 0.70 to 0.80). Calibration plots indicated that the model accurately predicted the risk of PMV (see Figure S5).

### Clinical utility

Based on the decision curve analysis in derivation cohort (see Figure 3A), when threshold probability was in the range of 0.10 to 0.77, using the two PAIRS scores to predict PMV would provide more net benefit than either a “treat-all-patients scheme” or a “treat-none scheme”. Moreover, the radiographic PARIS score had the highest net benefit when threshold probability was in the range of 0.10 to 0.84. These findings were consistent in the validation cohort as well (see Figure 3B).

**Figure 3.**
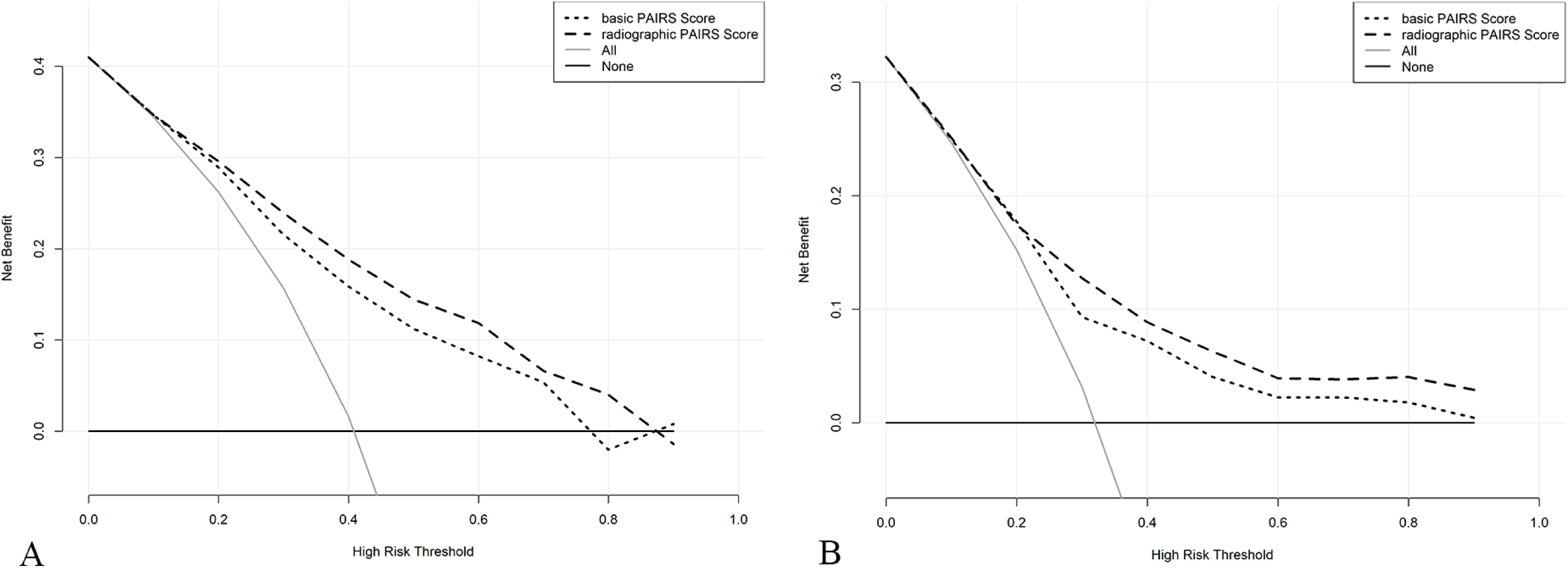
Decision curve analysis demonstrating the net benefit associated with the use of two PAIRS scores for predicting prolonged mechanical ventilation in derivation cohort (A) and in validation cohort (B).

### Comparison of two PAIRS scores

In the derivation cohort, adding radiographic results at follow-up improved the predictive performance of the model compared to the basic PAIRS score. This was evidenced by statistically significant increases in both the NRI (0.62, 95% CI 0.47 to 0.77, *P*<0.001) and IDI (0.05, 95% CI 0.03 to 0.07, *P*<0.001) for the radiographic PAIRS score.

Similar results were observed in the validation cohort, where the inclusion of radiographic results at follow-up also led to statistically significant improvements in NRI (0.55, 95% CI 0.38 to 0.72, *P*<0.001) and IDI (0.06, 95% CI 0.04 to 0.09, *P*<0.001).

## DISCUSSION

This investigation is pioneering in its exploration of the prevalence, predictive factors, and consequences of PMV in VBAO patients post-EVT. Our findings disclose a concerning high incidence of PMV among VBAO patients, even in the context of contemporary EVT techniques. This trend is alarmingly correlated with dire outcomes, including less than 10% favorable outcomes and an approximate 70% mortality rate at the 3-month mark. These insights underscore the critical need for a predictive model for PMV, which would significantly enhance clinical decision-making, facilitate communication with patients’ relatives, and improve the management of critically ill VBAO patients.

In our study, eight key variables were identified as potential predictors: reperfusion status, baseline NIHSS scores, collateral circulation status, history of AF, admission FBG levels, circulation insufficiency, and the presence of MCE and sICH in follow-up imaging. Notably, except for the two radiographic results which necessitate departmental scanning, the remaining six variables are readily obtainable at the ward post-EVT.

These identified predictors were integrated to formulate two versions of the PAIRS score, differentiated by the inclusion of the two imaging variables obtained at follow-up. Both versions of the PAIRS score demonstrated robust discriminative capabilities and model fit, as evidenced by internal validation using 1000 bootstrap resamples and external validation processes.

### Mechanism of associations

In patients with VBAO who require early PMV following EVT, defined as MV initiated within 7 days of VBAO onset, there is an indication of significant impairment in brain regions responsible for consciousness, breathing, and circulation. This often leads to catastrophic outcomes. Successful reperfusion has been identified as a critical predictor of favorable outcomes in VBAO patients.^22^ Achieving successful reperfusion can salvage a substantial volume of cerebral tissue in the posterior circulation, thereby mitigating damage to vital centers in the brainstem.

A higher NIHSS score correlates with an increased risk of stroke-associated pneumonia and, subsequently, a heightened risk of respiratory failure.^23^ Notably, components of the NIHSS score exceeding 30 in the PAIRS score system, indicative of severely impaired consciousness, are associated with an increased need for tracheostomy or a higher rate of extubation failure.^24,25^

Collateral circulation status, as evaluated by BATMAN score, plays a pivotal role in the pathophysiology of cerebral ischemia. An unfavorable BATMAN score (<7 points) significantly correlates with a poor outcome,^14^ possibly due to accelerated recruitment of the ischemic penumbra into infarction, resulting in extensive infarct size affecting respiratory or circulatory centers in the brainstem.

Interestingly, a history of AF emerged as a protective factor against PMV in VBAO patients post-EVT. This may be attributed to the cardioembolic mechanism often underlying VBAO in patients with AF, which is linked to higher rates of successful recanalization compared to atherosclerotic mechanisms.^26,27^ Consequently, this might mitigate damage to critical brainstem centers.

Our prior research has demonstrated that elevated admission FBG levels are significantly associated with an increased risk of poor outcomes and sICH.^28^ Hyperglycemia may exacerbate oxidative stress, compromise blood-brain barrier integrity post-cerebral ischemia/reperfusion injury, and heighten the risk of brain edema and hemorrhagic transformation, leading to expanded infarct volume and greater neurological deficits.^28^

Circulation insufficiency emerged as the strongest predictor for PMV in the PAIRS score. Patients with circulation insufficiency often struggle to meet the weaning criteria due to unstable cardiovascular status. Additionally, circulation insufficiency may signify impairment of the cardiovascular regulating center in the medulla, potentially caused by infarction or intracranial hypertension, thereby increasing the risk of cardiac arrest.^1^

Finally, the presence of MCE and sICH contributes to obstructive hydrocephalus and cerebellar tonsillar herniation. These conditions can induce secondary injury to the brainstem due to mass effect, thereby elevating the risk of impaired consciousness and failure of respiratory or circulatory functions, both of which are radiographically linked to PMV.^24^

### Clinical implications

In this study, we developed and validated two distinct versions of the PAIRS score, differentiated based on the inclusion of radiographic results at follow-up. Recognizing the challenges posed by transporting critically ill patients to medical imaging departments for CT or MRI scans, the basic PAIRS score was designed for practical application in such scenarios. In comparison, the radiographic PAIRS score, incorporating follow-up imaging data, demonstrated superior performance in risk reclassification and discrimination, as evidenced by positive NRI and IDI values. Decision curve analysis affirmed that both versions of the PAIRS score conferred substantial net benefits in predicting PMV across a wide range of thresholds, with the radiographic PAIRS score exhibiting the highest net benefit.

The PAIRS score serves as a valuable clinical tool, facilitating the stratification of patients into distinct risk categories for PMV. It assists clinicians in making informed decisions, communicating effectively with patients’ relatives, and optimizing patient care. Low-risk patients can be efficiently transitioned out of the ICU to conserve resources. Given the grave risk of catastrophic outcomes in PMV patients, considering palliative care for those at very high risk may be a judicious approach, despite the availability of intensive treatments.

Notably, the PAIRS score includes two modifiable factors — reperfusion status and blood glucose levels — offering potential targets for intervention in high-risk patients. Enhancing recanalization rates may further mitigate the incidence of PMV, and rigorous management of blood glucose levels is also crucial.

Our study has several limitations. Firstly, we lacked data on certain factors that could affect PMV, such as sepsis, heart failure, and sedation use, despite our efforts to include various study parameters. Secondly, the presence of missing data in our dataset might introduce bias in both the development and validation of our model. Lastly, the role of palliative care in influencing the outcomes of patients requiring PMV warrants consideration. For instance, VBAO patients post-EVT with sICH and MCE, rarely underwent suboccipital decompressive craniectomy. The potential efficacy of this surgical intervention in ameliorating catastrophic outcomes remains unclear.

## CONCLUSIONS

In this study, we identified factors associated with PMV in VABO patients, which can be assessed after EVT, and propose the two PAIRS scores to estimate the likelihood of PMV. These findings can assist patient families and caregivers in better understanding the treatment trajectory. Further validation of the effectiveness of the PAIRS score is necessary.

## Acknowledgments

None

